# Model training periods impact estimation of COVID-19 incidence from wastewater viral loads

**DOI:** 10.1101/2022.07.16.22276772

**Authors:** Maria L. Daza–Torres, J. Cricelio Montesinos-López, Minji Kim, Rachel Olson, C. Winston Bess, Lezlie Rueda, Mirjana Susa, Linnea Tucker, Yury E. García, Alec J. Schmidt, Colleen Naughton, Brad H. Pollock, Karen Shapiro, Miriam Nuño, Heather N. Bischel

## Abstract

**Background:** Wastewater-based epidemiology (WBE) has been deployed broadly as an early warning tool for emerging COVID-19 outbreaks. WBE can inform targeted interventions and identify communities with high transmission, enabling quick and effective response. As wastewater becomes an increasingly important indicator for COVID-19 transmission, more robust methods and metrics are needed to guide public health decision making.

**Objectives:** The aim of this research was to develop and implement a mathematical framework to infer incident cases of COVID-19 from SARS-CoV-2 levels measured in wastewater. We propose a classification scheme to assess the adequacy of model training periods based on clinical testing rates and assess the sensitivity of model predictions to training periods.

**Methods:** We present a Bayesian deconvolution method and linear regression to estimate COVID-19 cases from wastewater data. We described an approach to characterize adequacy in testing during specific time periods and provided evidence to highlight the importance of model training periods on the projection of cases. We estimated the effective reproductive number (*R*_*e*_) directly from observed cases and from the reconstructed incidence of cases from wastewater. The proposed modeling framework was applied to three Northern California communities served by distinct wastewater treatment plants.

**Results:** Both deconvolution and linear regression models consistently projected robust estimates of prevalent cases and *R*_*e*_ from wastewater influent samples when assuming training periods with adequate testing. Case estimates from models that used poorer-quality training periods consistently underestimated observed cases.

**Discussion:** Wastewater surveillance data requires robust statistical modeling methods to provide actionable insight for public health decision-making. We propose and validate a modeling framework that can provide estimates of COVID-19 cases and *R*_*e*_ from wastewater data that can be used as tool for disease surveillance including quality assessment for potential training data.

## 1 Introduction

During the early phases of the COVID-19 pandemic, caused by the severe acute respiratory syndrome coronavirus 2 (SARS-CoV-2), the World Health Organization (WHO) recommended implementing mass testing programs as a containment measure. Individual diagnostic testing informs contact tracing and medical interventions, ideally cutting chains of transmission short and containing outbreaks. Mass clinical screening programs can also provide valuable data on community-level health trends, but maintaining mass testing programs for the purpose of community-level monitoring is expensive and requires robust infrastructure with consistent availability of testing supplies and human resources. ^1^ Moreover, diagnostic tests validated in low-throughput clinical settings (like nucleic acid amplification tests or NAATs) are not necessarily efficient platforms for constructing community screening programs. ^2^ Design-wise, employing such tests for large-scale screening requires extensive logistical coordination over large geographic areas. This becomes especially complicated when the options for diagnostic tests are myriad, lack standardization, and depend heavily on local social landscapes. Small biases in the tests may be inflated when deployed broadly, leading to large spurious associations at the population level. ^3,4^

Public health authorities are turning to wastewater-based epidemiology (WBE) as an alternative strategy for unbiased population-level surveillance of COVID-19. WBE uses biomarkers in wastewater to monitor trends in community-level health indices. WBE methods have been used to detect changes in drug consumption, ^5,6^ dietary patterns, ^7^ and the circulation of pathogens like poliovirus and norovirus. ^8^ Measurements of SARS-CoV-2 RNA in wastewater correlate strongly with changes in COVID-19 prevalence in the associated communities. ^9–12^ Since the onset of the pandemic, WBE of SARS-CoV-2 has been implemented in over 67 countries and 279 universities. ^13^ In some places, WBE programs have detected changes in SARS-CoV-2 RNA levels in wastewater prior to changes in local COVID-19 hospitalization activity and spikes in NAAT screening cases. ^14–19^ Others have used WBE to assess the effectiveness of public health interventions, ^20^ and recently, WBE was used to predict hospitalizations and ICU admissions. ^21^ In addition to monitoring trends, WBE can provide estimates of critical disease transmission parameters in the community without the biases associated with test-seeking behavior or poor access to testing programs.

An ongoing challenge for WBE is developing robust data collection and interpretation methods that are comparable across time and geography. Variation in sampling design and sample processing methods, natural variability in viral shedding rates in feces, variability in wastewater flow volume, population fluctuations, and location-specific characteristics of wastewater management are all factors that make inference of new COVID-19 cases from wastewater data challenging. ^22–25^ Such factors will ultimately affect uncertainty estimates when modeling disease incidence and other public health indicators. An ideal WBE program would implement a generalized approach that provides consistent estimates of disease burden in a targeted population, yielding pubic health metrics like the disease incidence, disease prevalence and/or the effective reproductive number (*R*_*e*_). Previous studies that approach this problem include: simple algebraic adjustments with environmental constants ^26^; estimating the total number of cases with a susceptible-exposed-infectious-recovered (SEIR) model informed by wastewater results^11^; using regression analysis to estimate the number of infected people ^9^; and making near real-time estimates of *R*_*e*_. ^10,27^

We propose and compare two modeling approaches: a simple linear model and a Bayesian deconvolution approach to estimate COVID-19 incident cases from wastewater viral loads. Both models rely on short training periods to calibrate wastewater measurements using clinical testing data from a community screening program. We evaluate the impact of different training periods on model predictions, hypothesizing that relative rates of change in clinical testing and reported cases can be used to identify appropriate model training periods. We then apply the framework to estimate incident cases and *R*_*e*_ from wastewater influent data generated for three communities in Northern California. The methodology we describe can be generalized to other WBE systems to track the evolution and assess the magnitude of COVID-19 fluctuations and outbreaks in a manner that is comparable across programs, locations, and time.

## 2 Methods

The analytical framework was developed using data from the City of Davis (Davis) and replicated for the City of Woodland and the University of California Davis. Analysis includes case and wastewater (WW) data from December 1, 2021 to March 31, 2022. Results for the City of Woodland and UC Davis are presented in the Supplementary Materials section.

### 2.1 Wastewater sample collection

Staff from three Northern California wastewater treatment facilities (Davis, Woodland, and UC Davis) provided 24-hour composite wastewater samples 5-7 days per week. Samples were acquired using Teledyne ISCO 5800 refrigerated autosamplers in Davis and Woodland and a Hach Sigma 900 autosampler for UC Davis. The autosampler in Davis was programmed to collect 400 mL of influent every 15 “pulses”, where one pulse was set at 10,000 gallons. An average of 24 pulses was expected per day based on an average daily influent flow of 3.6 million gallons per day (MGD). The autosampler in Woodland was programmed to acquire 100 mL of influent every 15 min over a 24-hours period. The autosampler for UC Davis was programmed to acquire approximately 200 mL of influent every 20 min over a 24-hours period. The reported sample collection date corresponds to the date when an autosampler program was completed. Davis and UC Davis provided 12 ml samples in new 15-ml polypropylene centrifuge tubes. Woodland provided 1 L samples in Nalgene bottles that were washed, sterilized, and reused over the duration of sampling. Samples were stored at 4°C and transported weekly in coolers on ice to the analytical lab at UC Davis. For biosafety compliance, samples were placed in a water bath set at 60°C for 30 minutes and returned to 4°C prior to sample processing. Concentration and extraction were performed in a biosafety level 2 (BSL2)-certified laboratory.

### 2.2 Sample concentration and extraction

The sample concentration and extraction protocol were adapted from Karthikeyan et al. ^28^ using 4.875 mL instead of 10 mL starting sample volume. Each wastewater sample was deposited into a separate well of a KingFisher 24 deep-well plate (Thermo Fisher). An extraction control blank (nuclease-free water) was included in 90% of the deep-well plates to assess potential contamination during concentration and extraction. Each well was spiked with 50 *µ*L of Nanotrap^®^ Enhancement Reagent 1 (Ceres Nanosciences product ER1 SKU # 10111-10, 10111-30) and 5 *µ*L of a stock of vaccine-strain Bovine Coronavirus (BCoV, Bovilis^®^ Coronavirus vaccine) containing an estimated 1.3*x*10^8^ gc/mL as measured by ddPCR. 500 *µ*L aliquots of the initial BCoV vaccine stock, prepared from the suspension of lyophilized BCoV vaccine in 20 mL buffer provided with the kit, were stored at −80°*C* prior to use. Each spiked sample was manually agitated by pipetting up and down at least three times using a 5 mL pipette. Samples were then incubated for 30 minutes at room temperature. Following incubation, concentration was carried out using 75 *µ*L Nanotrap^®^ Magnetic Virus Particles (Ceres Nanosciences) on a KingFisher Apex robot (Thermo Scientific). Concentrated viruses were eluted from the Nanotrap^®^ beads into 400 mL of lysis buffer per sample from the MagMAX Microbiome Ultra Nucleic Acid Isolation Kit (Thermo Fisher). Concentrated samples were extracted per the MagMAX kit manufacturer instructions in 96 deep-well plates on the KingFisher Apex. Samples were eluted in 100 *µ*L of MagMAX Elution Solution. Extracts were typically stored on ice and immediately subjected to same-day analysis. When same-day analysis was not possible, extracts were immediately stored at −80°*C* until analysis.

### 2.3 Extract analysis by ddPCR

Sample extracts were analyzed by digital droplet polymerase chain reaction (ddPCR) for four targets: N1 and N2 targeting regions of the nucleocapsid (N) gene of SARS-CoV-2, and Bovine Coronavirus (BCoV) and pepper mild mottle virus (PMMoV) for normalization of the SARS-CoV-2 results. N1/N2 and BCoV/PMMoV were quantified in separate duplex assays. Due to high levels of PMMoV, the sample for the PMMoV/BCoV duplex was diluted 40x prior to loading. The duplex ddPCR amplifications were performed in 20 *µ*L reactions on a QX ONE ddPCR System (Bio-Rad). Each reaction contained the following components: 1x Supermix, 20 *U/µ*L Reverse transcriptase, 15 mM Dithiothreitol from the One-Step RT-ddPCR Advanced Kit for Probe (Bio-Rad), 900 nM of each primer, 250 nM of each probe, and 5 *µ*L of sample extract or control. The one-step ddPCR reaction consisted of 3 min plate equilibrium at 25°*C*, 60 min reverse transcription at 50°*C*, 10 min enzyme activation at 95°*C*, followed by 40 cycles of 30 s denaturation at 94°*C* and 1 min annealing/extension at 58°*C*, and then 10 min enzyme deactivation at 98°*C* and 1 min droplet stabilization at 25°*C*. Preparation and plating of ddPCR master mix were carried out in a separate location from sample loading to avoid contamination. Sample loading was performed using an epMotion ^®^ 5075 (Eppendorf) liquid handler. Each ddPCR plate included duplicate positive controls (stock mixture of synthesized gene fragments containing for the four target regions) for each target and duplicated no-template controls (nuclease free water). Additional information on the ddPCR assay designs is available in Tables S1–S4. Table S1 summarizes primers, probes for ddPCR assays performed as part of this work. Table S2 and S3 provide the ddPCR reaction and 20X primer/probe mix recipes. From October 21 to December 21, Cy5 and Cy5.5 were used in place of FAM and HEX as the fluorophores for PMMoV and BCoV, respectively. Table S4 lists details for the positive controls. Prior to 12/21/22 the annealing temperature was 60°*C*. The selection of positive and negative droplet clusters in samples and controls was conducted manually based on visual inspection of clusters. Results were considered invalid if the distribution of positive or negative droplets appeared abnormal in shape or if the total number of droplets generated fell below a threshold of 10,000 droplets in a single well. Additional information on data processing and quality control is provided in the Supplemental Material. We utilize N/PMMoV (the average SARS-CoV-2 RNA concentration (N) divided by the concentration of PMMoV) as the resulting WW signal for subsequent model development.

### 2.4 COVID-19 case data

Healthy Davis Together (HDT) and Healthy Yolo Together (HYT) provided daily COVID-19 cases and total tests performed during the study period for the City of Davis, UC Davis, and Woodland,from the community screening program. ^29,30^

Data were smoothed for the implementation of the linear model using a 7-day moving average (the mean of the current and the previous six days). This approach improves harmonization between the current WW concentration and observed cases (Figure 1B). The 7-day moving average of cases for the linear model is similar to a deconvolution model with equal weights (uniform shedding load distribution) and a shedding time of 7 days (Section 2.6.1). Rate of change for tests administered and positive cases were calculated from a weekly aggregation of daily test counts and positive cases identified. Changes in test and case rates were then used to determine training periods with adequate testing.

**Figure 1:**
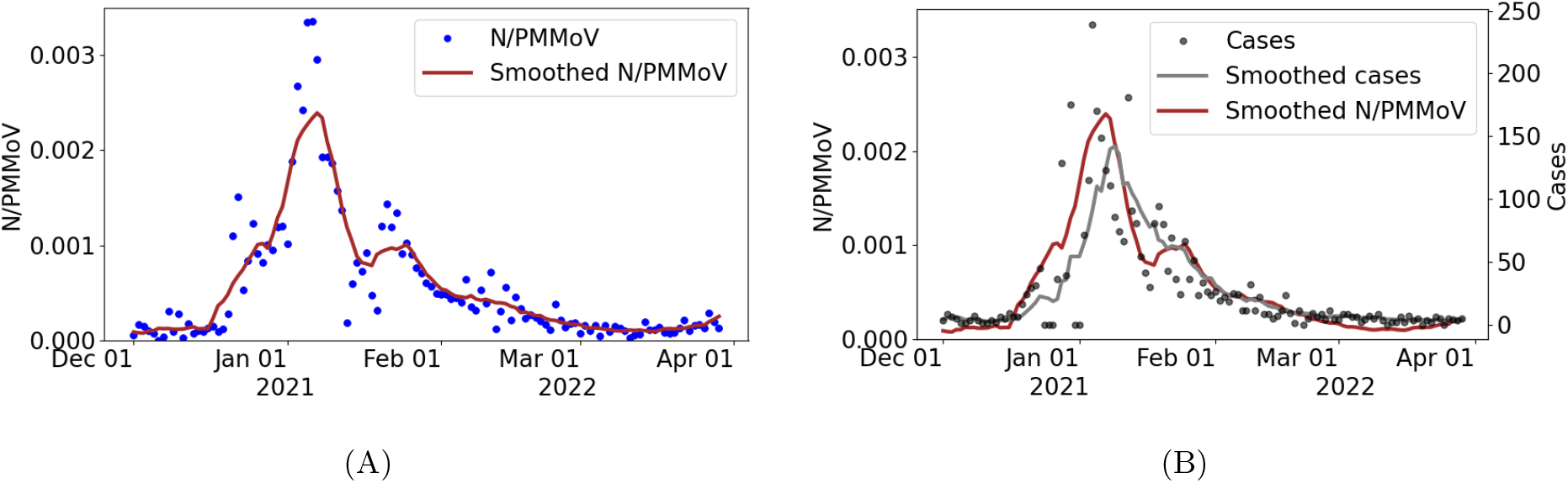
(A) Wastewater data (N/PMMoV) and 10-day moving average (Smoothed N/PMMoV) from December 1, 2021 to March 31, 2022. (B) Cases, 7-day moving average smoothed cases, and 10-day moving average of smoothed WW data.

### 2.5 Smoothed wastewater signal

To reduce uncertainty and to minimize daily fluctuations of cases observed, we applied a 10-day moving average for daily influent WW data (Figure 1A). We use the resulting smoothed influent WW data to correlate with smoothed cases (Figure 1B).

### 2.6 Models

We present two models to estimate COVID-19 cases from SARS-CoV-2 RNA in the WW. The first model was adapted from Huisman et al.’s approach ^10^ and relates past infections with WW signal through the convolution described in Equation (3). The number of daily cases is modeled with a negative binomial (NB) distribution through the deconvolution (the inverse operation of convolution) noted in Equation (3). The second approach uses a simple linear regression to estimate cases (dependent variable) from WW data (independent variable). We also propose a strategy for selecting model training periods with adequate clinical testing to estimate parameters and improve estimation.

#### 2.6.1 Deconvolution model

Viral RNA concentrations measured in wastewater (*C*_*i*_’s) are related to the number of new infections per day (*I*_*i*_’s) through the profile of SARS-CoV-2 RNA shedding in the wastewater by an infected individual days after infection or symptom onset. ^10^ The measurement *C*_*i*_ of WW on day *i* is related to infections *I*_*j*_ on prior day *j* through the following convolution:

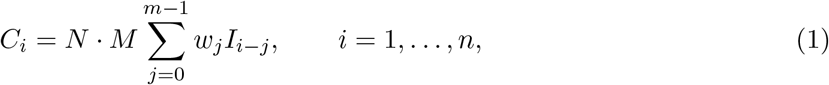

where *w*_*j*_, *j* = 1, …, *m* (sums to 1) is the shedding load distribution describing the temporal dynamics of shedding and *m* is the duration of viral shedding or shedding time. The normalization factor *N* represents the total virus shed by an infected individual during the infection period. *M* is a constant that depends on the sewer system, wastewater treatment plant, and processing pipeline. The measurement of viral RNA in wastewater *C*_*i*_ on the day *i* is used to estimate COVID-19 cases from wastewater concentration data via convolution. As noted by Huisman et al., ^10^ normalization factors *N* and *M* are difficult to measure, and they assume *B* = *N* · *M* as the lowest concentration of the viral load or concentration from a single infection.^10^ The weights for shedding load distribution (*w*_*j*_) can be estimated using individual-pooled analysis of SARS-CoV-2 viral loads. ^31–33^ Instead, we estimate *B* and corresponding weights using measured WW data and cases within a specified period (training period) by directly modeling the deconvolution process through a Bayesian approach.

We model *w*_*j*_ as follows:

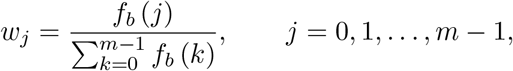

where *f*_*b*_ (*k*) is the probability density function (pdf) of a random variable *X* with exponential distribution of rate parameter *b*. Hereafter, notation 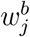 will be used instead of *w*_*j*_ emphasizing that weights depends on parameter *b*. Notice that, if *X* ∼ *Exp*(*b*) then its pdf is *f*_*b*_ (*k*) = *be*^*−b·k*^, thus:

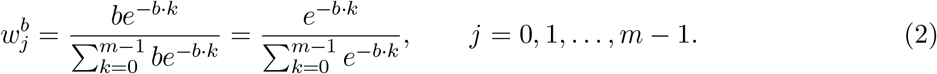

Equation (1) is rewritten using Equation (2) as follows:

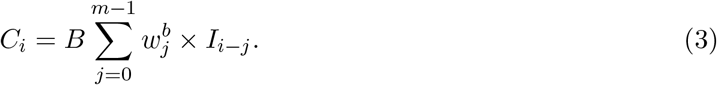

The deconvolution of the Equation (3) will be denoted as *dec*(**C**, *B, b*), where **C** = (*C*_1_, *C*_2_, … *C*_*n*_) represents the vector of WW data and parameters *B* and *b* are described above. **I** = (*I*_1_, *I*_2_, …, *I*_*n*_) correspond to daily cases counts and its theoretical expectation is given by (𝔼(**I**) = ***µ***) estimated in terms of the deconvolution model as ***µ*** = (*µ*_1_, *µ*_2_, …, *µ*_*n*_) = *dec*(**C**, *B, b*). The deconvolution is approximated using the Richardson–Lucy algorithm. ^34^

##### Observational model

We estimate the number of COVID-19 cases per day (*I*_*i*_’s) using the Negative Binomial (NB) model, which is most relevant for overdispersed count data. In this situation, the variance exceeds the mean. The NB distribution describes a sequence of independent and identically distributed Bernoulli trials with a probability of success *p* before a specified (non-random) number of successes (*r*) occurs. Assuming a similar approach as in Lindén et al., ^35^ we reparametrized the NB distribution in terms of its mean *µ* and “overdispersion” parameters *ω* and *α*, with 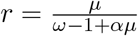 and 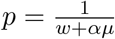 in the usual NB parametrization. We assume that *I*_*i*_ follows a NB distribution. Denoting the mean and variance as *µ*_*i*_ and 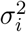, respectively, and requiring that 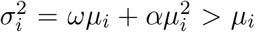, we enforce overdispersion for suitable chosen parameters *ω* and *α*. The index of dispersion is 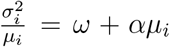. Overdispersion concerning the Poisson distribution is achieved when *ω >* 1 and the index of dispersion increases with size if *α* ≠ 0; adding variability as counts increase. We found good performance fixing *ω* = 2 and *α* = 0.05, implying higher variability for the later.

Using the deconvolution model and parameter as described above, we obtain the following likelihood function with the assumed NB model:

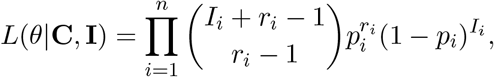

where 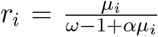 is the number of successes, 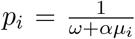 is the probability of a single success, and (*µ*_1_, *µ*_2_, …, *µ*_*n*_) = *dec*(**C**, *B, b*).

We estimate *θ* = (*B, b*) from measurements of WW data **C** = (*C*_1_, *C*_2_, … *C*_*n*_) and COVID-19 cases **I** = (*I*_1_, *I*_2_, … *I*_*n*_). We adopt a Bayesian statistical approach, which is well suited to model multiple sources of uncertainty and allows the incorporation of background knowledge on the model’s parameters. In this framework, a prior distribution, *π*_Θ_(*θ*), is required to account for unknown parameter *θ* in order to obtain the posterior distribution. For *b*, we assumed a Gamma distribution with shape and scale parameters *v*_*b*_ = 2 and *S*_*b*_ = 1, respectively; this assumption is based on published data on viral shedding duration in gastrointestinal samples. ^36^ For *B*, we assumed a Gamma distribution with shape and scale parameters *v*_*M*_ = 2 and *S*_*M*_ = 2*/*1*e*^*−*4^, respectively; based on the lowest viral RNA concentrations observed. Having specified the likelihood and the prior, we use Bayes’ rule to calculate the posterior distribution,

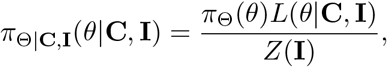

where *Z*(**I**) = ∫ *π*_Θ_(*θ*)*L*(*θ*|**C, I**)*dθ* is the normalization constant. The posterior distribution is simulated using a t-walk Markov chain Monte Carlo (MCMC) algorithm. ^37^

##### Duration of viral shedding

The deviance information criterion (DIC) was used to select the shedding time (*m*). DIC is a Bayesian generalization of the Akaike information criterion (AIC) for model selection in a finite set of models, with preference given to models with lower DIC. The DIC is preferred in settings with Bayesian model selection problems where the model’s posterior distributions are obtained by MCMC simulation. ^38^ We selected the appropriate shedding time by computing DIC in a grid search along the parameter space *m* : {6, …, 10}.

##### Simple linear regression model

We assume the following noise model,

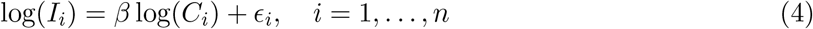

where *I*_*i*_ is the number of new infections on day *i, C*_*i*_ is the measurement of viral RNA in the wastewater on the *i*th day, and *ϵ*_*i*_ is a random residual associated with day *i* which is assumed to be distributed as *N* (0, *σ*^2^), with *σ*^2^ as the residual variance. This inference problem aims to estimate *θ* = (*β, σ*) from WW data and cases. A log-linear model is assumed to address positively skewed data and prevent negative fitted values.

### 2.7 Selection of model training periods

We describe whether or not testing is adequate in a particular period of observed cases by calculating the rate of change in tests conducted and new cases within a specific time period. We define the rate of change of both tests conducted 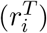 and confirmed positive cases 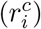 during period *i* as 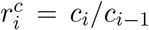 and 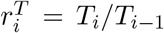, respectively, where *T*_*i−*1_, *T*_*i*_ denote the number of tests carried out in two consecutive periods, and *c*_*i−*1_, *c*_*i*_ denote the number of positive cases detected in these periods. We classify a testing period as adequate when the rate of change in testing is greater than the rate of change in cases; otherwise, if the rate of change in testing is lower/equal to the rate of change in cases, we conclude that testing is inadequate. We summarize various scenarios of testing adequacy in Table 1. Our determination of testing adequacy, and thus suitability for model training for both linear and deconvolution models, assumes that observed cases would be sufficiently close to actual cases when testing rates are high compared to case rates and test positivity remains low as determined through the community screening programs.

**Table 1:** Test and case scenarios to assess adequacy in testing for training periods.

| Scenario | Sub-Scenario | Classification |
| --- | --- | --- |
| Testing and cases increase, $T_i \geq T_{i-1}$, $c_i \geq c_{i-1}$ | Cases increase faster than testing, then $r_i^T \leq r_i^c$ | Not adequate |
| | Testings increase faster than cases, then $r_i^T \geq r_i^c$ | Adequate |
| Testing and cases decrease, $T_i < T_{i-1}$, $c_i < c_{i-1}$ | Testing decreases faster than cases, then $r_i^T < r_i^c$ | Not adequate |
| | Cases decrease faster than tests, then $r_i^T > r_i^c$ | Adequate |
| Testing increase and cases decrease, $T_i \geq T_{i-1}$, $c_i < c_{i-1}$ | Then, $r_i^T \geq r_i^c$ | Adequate |
| Testing decrease and cases increase, $T_i < T_{i-1}$, $c_i \geq c_{i-1}$ | Then, $r_i^T \leq r_i^c$ | Not adequate |

### 2.8 Effective reproductive number

The number of people in a population who are susceptible to infection by an infected individual at any particular time is denoted by *R*_*e*_, the effective reproductive number. This dimensionless quantity is sensitive to time-dependent variation due to reductions in susceptible individuals, changes in population immunity, and other factors. *R*_*e*_ can be estimated by the ratio of the number of new infections (*I*_*t*_) generated at time *t*, to the total infectious individuals at time *t*, given by 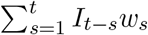, the sum of infection incidence up to time step *t* − 1, weighted by the infectivity function *w*_*s*_. We implemented Cori et al.’s approach ^39^ to estimate *R*_*e*_ directly from observed cases and from cases that were estimated from the WW data.

## 3 Results

### 3.1 Identification of adequate training periods

We computed the rate of change in the number of tests and cases by week for the City of Davis between December 1, 2021, and March 31, 2022 (Figure 2 and Table S6). Each week was compared with a previous week and classified as adequate whenever the rate of change in tests was greater than the rate of change in cases and as not adequate otherwise.

**Figure 2:**
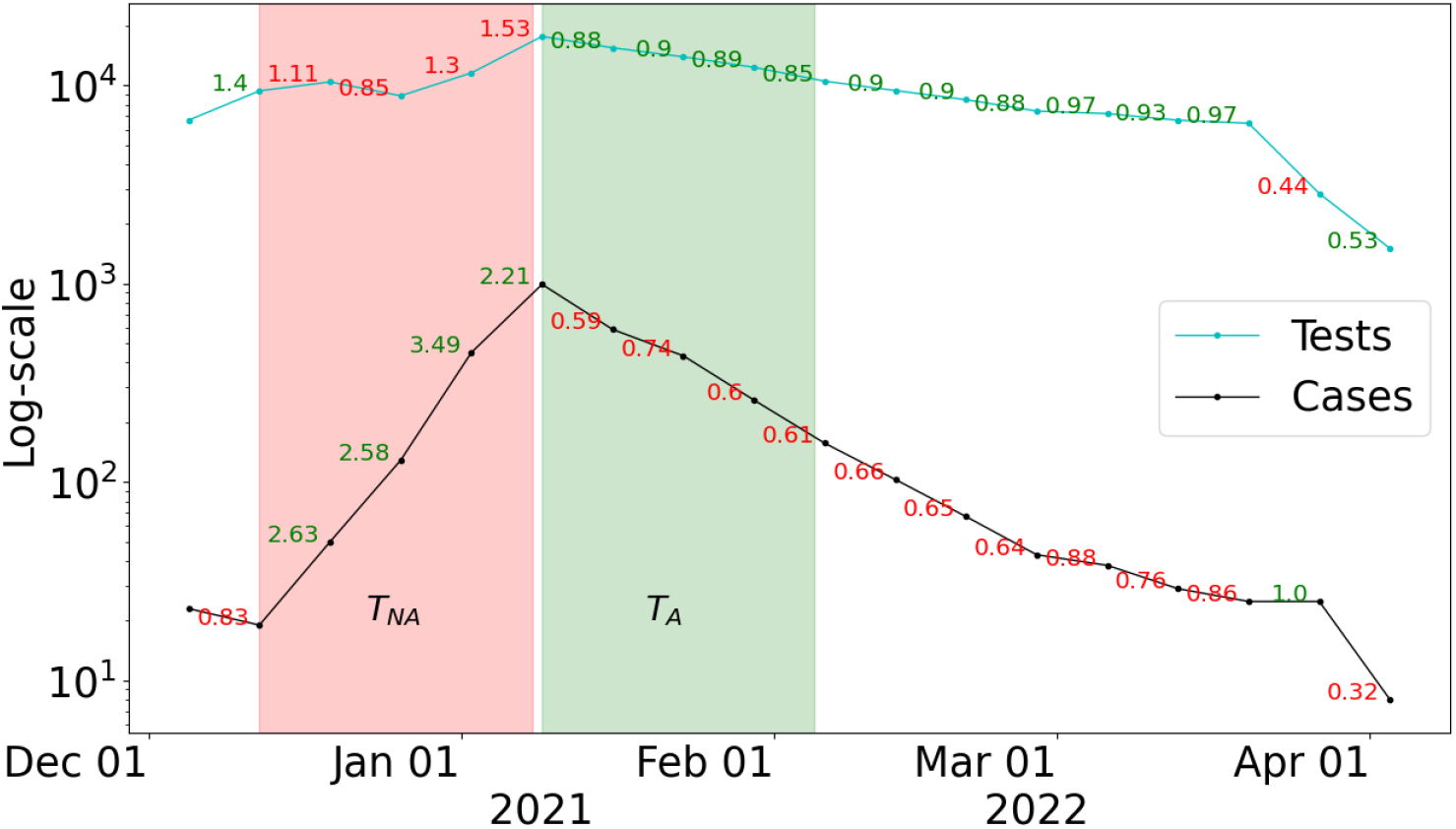
Number of tests administered in the City of Davis (cyan line) and cases (black line) by week, on a log-scale, from December 1, 2021 to March 31, 2022. The week-to-week rate of change in cases and tests are displayed; green numbers indicate the test rate is greater than the case rate, and red numbers are the opposite. The green and red shaded region correspond training periods with Adequate (*T*_*A*_) and Not Adequate (*T*_*NA*_) testing, respectively.

Figure 2 illustrates two specific training periods assumed for the analysis of the City of Davis. The first training period includes data from December 12, 2021 to January 8, 2022 (red region, denoted by *T*_*NA*_), and the second training period assumes data from January 9 to February 2, 2022 (green region, *T*_*A*_). A training period designated by *T*_*NA*_ (Not Adequate) corresponds to a scenario where test rate is consistently lower than the rate of new cases. Similarly, a training period denoted by *T*_*A*_ corresponds to a scenario in which the testing rate exceeds the rate of new cases. We assess testing adequacy for Woodland and UC Davis (Figures S1 and S3), and similarly identified a period of inadequate testing prior to an observed surge in infections..

### 3.2 Comparison of models to estimate public health metrics from wastewater data

We applied both a deconvolution technique and a linear regression to reconstruct incident cases of COVID-19 from the WW data, assuming model training periods according to the adequacy of clinical testing efforts. We found that the magnitude of case projections and trends was sensitive to the assumption of the model training period for both model constructs (Figure 3), but the timing of peaks in cases predicted were independent of the training period. Case predictions from the models that assumed a training period with inadequate testing (*T*_*NA*_) were consistently lower than projections from the models that assumed a training period with adequate testing (*T*_*A*_). These results suggest that models using *T*_*NA*_ systematically underestimated true case counts, a finding consistent with our expectations since fewer cases are detected during periods of inadequate clinical testing than when testing is adequate. Projection of cases from the models using *T*_*A*_ aligned more consistently with observed cases in periods where testing was deemed adequate. The difference in predictions in cases from the two training periods was particularly evident in January 2021, during the onset of the Omicron variant surge in Davis.

**Figure 3:**
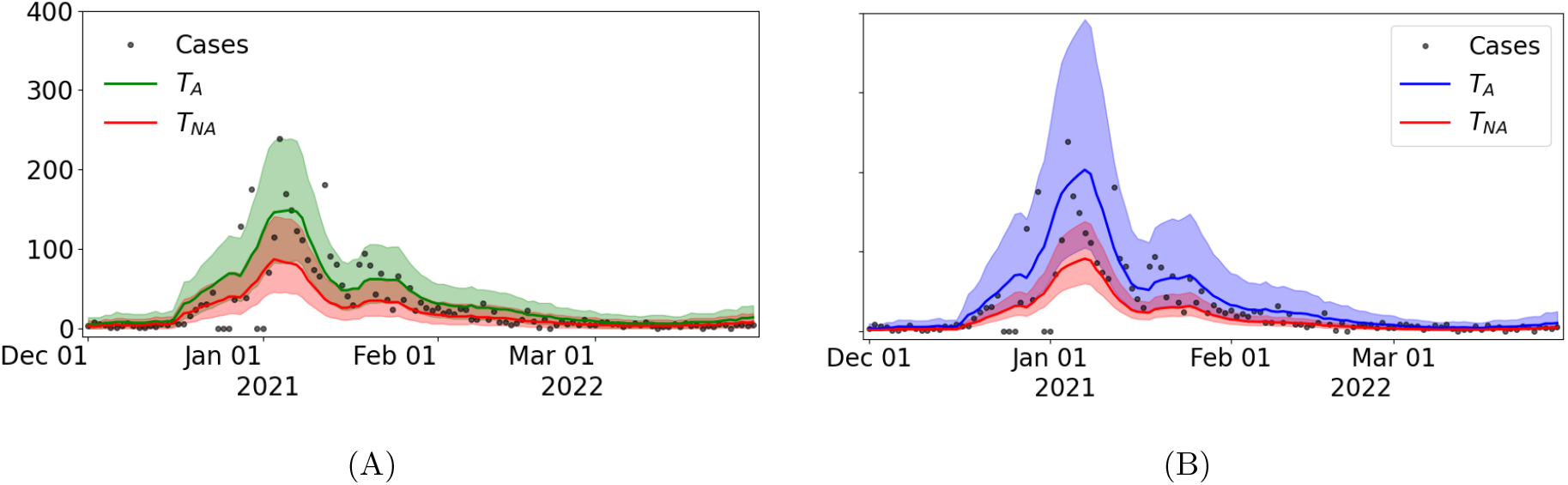
Predicted cases assuming (A) the deconvolution and (B) the linear regression models from WW data between December 1, 2021, and March 31, 2022. The estimated cases are displayed in green(blue) for the deconvolution(linear) model, when the models were trained on the period classified as adequate (*T*_*A*_), and in red when both models were trained on the period classified as inadequate (*T*_*NA*_). Solid lines represent median estimates of cases, and 95% prediction intervals are depicted in shaded regions.

Incident case projections from the linear model that assumed *T*_*A*_ was able to capture the peak of the curve more closely than the deconvolution model, although with greater uncertainty. It is worth noting that results from the deconvolution and the linear models are similar because the linear model is fitted with the 7-day moving-average of case data. Data smoothing of this kind corresponds to a convolution with equal daily weights. The estimation of cases from the linear regression model that *T*_*NA*_ was similar to the results of the deconvolution model, Figure 3B.

*R*_*e*_ monitors changes in disease transmission over time, assesses the effectiveness of interventions, and can be useful to guide policy decision making. Estimates of *R*_*e*_ from the median of the predicted cases using the deconvolution model and the linear model are similar. In most of the time period assessed, *R*_*e*_ determined from WW results are quite similar in magnitude and follow the trends for *R*_*e*_ calculated using observed cases (Figure 4B). A notable difference between the *R*_*e*_ estimated with the observed cases and that obtained with the WW data with both the linear model and the deconvolution model occurs at the end of March. At this time, the median of *R*_*e*_ for cases is below 1, and the median of *R*_*e*_ with WW data for the linear and deconvolution model are above 1. A value of *R*_*e*_ lower than 1 indicates that overall transmission is declining, while thresholds higher than 1 suggest that an outbreak is expected to continue. ^39^ The posterior analysis using the estimated *R*_*e*_ from WW data thus indicates that an outbreak may have occurred in Davis that was not detected through clinical cases Figure S5.

**Figure 4:**
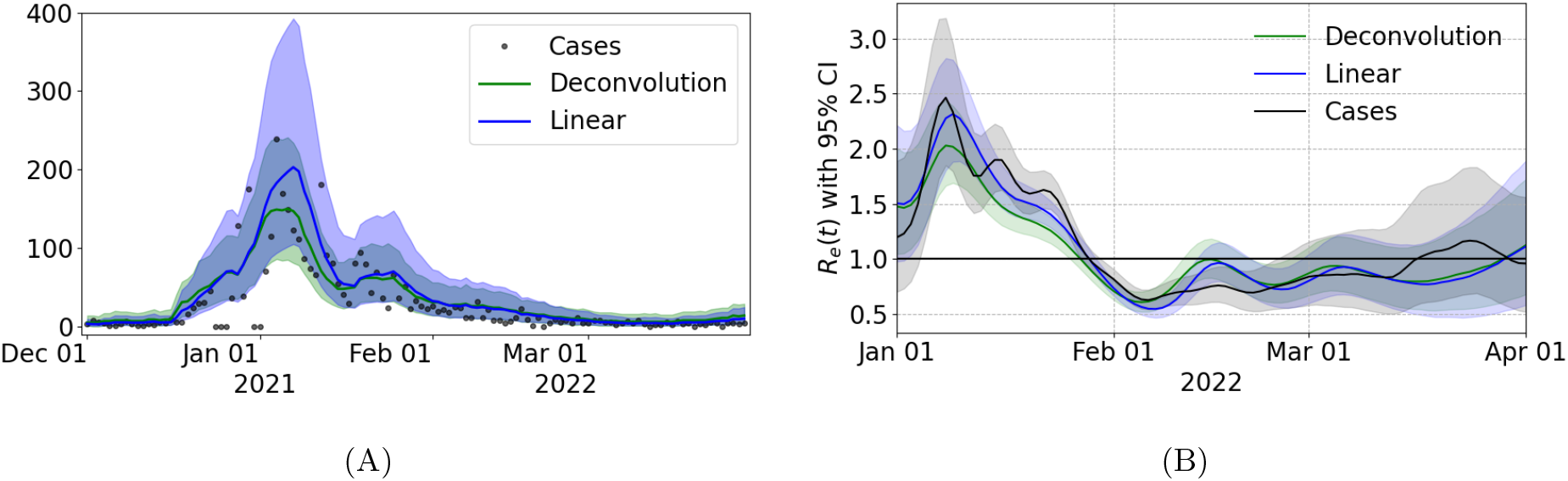
(A) Cases predicted with the linear regression model (blue) and the deconvolution model (green) using WW data between December 1, 2021, and March 31, 2022. Both models were trained in the period classified as appropriate (*T*_*A*_). (B) Effective *R*_*e*_ of city of Davis computed with the median of the cases estimated for the deconvolution model (blue) and the linear regression (green).

We demonstrate the adaptability of our methodology using data for Woodland and UC Davis and present results in the Supplemental Material (Figures S1 and S3). The trends of the observed cases is recovered with both models (Figures S2 and S4), yielding results consistent with those obtained for Davis.

The code implemented for the study are available in the github repository **??**. Analyses were carried out using Python version 3.

## 4 Discussion

Community-wide testing has played a critical role in mitigating the COVID-19 pandemic. However, large-scale testing has been limited and falls further behind during surges of infections. We developed criteria to classify the adequacy of clinical testing in a community through time, and we applied the classification scheme to three Northern California communities. As was observed in many other communities at the time, we found that clinical testing was inadequate at the front end of the wave of Omicron infections that occurred during our study period. Inadequate clinical testing during surges of infection makes it particularly challenging to discern true levels of SARS-CoV-2 infections in a population. WBE can fill data gaps caused by inadequate testing programs. As clinical testing transitions further towards at-home self-testing, measurements of SARS-CoV-2 RNA in wastewater can serve as an increasingly important indicator for COVID-19 transmission.

Myriad sources of variability and uncertainty in WW data can nevertheless impact the accuracy of estimates of COVID-19 cases or other public health metrics derived from WW data. ^40–43^ Statistically representative samples can also be difficult to obtain because of the complexity of wastewater collection systems and the physical challenge of ensuring consistency in sample acquisition and processing. ^44^ Such challenges can limit comparability of wastewater results across different WBE programs. The modeling framework we described to estimate COVID-19 cases from WW data accounts for uncertainty and relies on short training periods using clinical testing data to calibrate wastewater measurements to local conditions.

We showed that case projections reconstructed from either the Bayesian deconvolution or the simple linear model were generally higher than cases observed through clinical testing, particularly during periods with sub-optimal testing. These results are not surprising, as we expected that the WW models would yield case estimates higher than cases observed through screening given that WW is not subject to the same selection biases as testing. While both the deconvolution and linear regression models captured the overall trends in observed cases overall, qualitative differences were evident between the approaches, particularly when testing was limited. Both models identified steep upward trends in cases during the surge in mid-January and at the onset of the Omicron surge Figure 4A.

The classification approach that we developed to assess the adequacy of model training periods was essential to providing robust estimates of case projections. Training periods that satisfied the proposed characteristics (i.e., adequate testing) resulted in similar estimates from each of the models, and yielded trends that were consistent with observed cases. Case projections that assumed training periods with poor testing generally underestimated cases compared to projections from adequate training periods. While the proposed models do not seek to recover the curve of reported cases (an underestimate of actual cases specially in a limited testing scenario), use of adequate training periods for the WW models enabled us to capture trends in case counts much more closely. It is evident that training periods with inadequate testing introduce a downward bias into the model.

WBE has substantially lower resource requirements than mass diagnostic testing, and WW data lack bias from care- and test-seeking behavior in the catchment population. WBE programs that determine COVID-19 public health metrics at the community level can work as a powerful and cost-effective complement to other, more traditional intervention methods. The analytic methods presented here can inform local public health policy and community-level interventions, for instance by helping to assess when initiation of clinical screening programs and non-pharmaceutical interventions are needed. The model can be especially valuable to fill data gaps during surges of infection when clinical testing is inadequate and could be used to assess when estimates of case rates exceed certain thresholds. WBE does come with the inherent challenge of determining the populations being monitored, the effect of which is exacerbated if the population served is highly mobile (e.g., a university campus). In other words, WBE methods for tracking COVID-19 are inherently location-specific, whereas public screening programs are tied to the people they serve. Calibration of wastewater models using clinical data will be most robust in places with minimal mobility or within populations that are adequately described and understood.

With screening programs winding down across the United States, finding training periods with adequate testing rates may not be feasible. In such cases, application of the deconvolution model for WBE can still highlight important trends. Where tests are only administered for clinical diagnostics, a periodic sentinel system could be employed to produce sufficient prevalence estimates for training periods. Such a system would recruit a representative sample of the population for repeated testing during a training period to establish a baseline, which then enables the wastewater deconvolution model to track incidence for an extended period of time. The same sentinel group could be called back later when the model needs to be updated to retrain for new situations.

WBE surveillance systems should be cognizant that they are not unduly targeting and stigmatizing vulnerable communities. WBE is much less invasive than diagnostic testing and protects individual identities, thereby avoiding the stigmatization of individuals and not requiring individual consent.^45^ Yet focusing too heavily on public surveillance efforts can negatively influence public perception of those being monitored. ^46^ Mathematical models that employ machine learning, such as the deconvolution model described herein, must be trained with data sets that are not sampled by biased collection methods, else they may inadvertently reintroduce social biases into the results and contribute to larger inequities in public health.

## Data Availability

All data produced in the present study are available upon reasonable request to the authors.

## Human Participant Protection

This study was determined to be exempt from institutional review board review by the UC Davis Office of Research.

## Acknowledments

M.L.D.T and J.C.M.L., literature review, model development, implementation, wrote overall first draft of paper, paper revisions. M.K., analytical assay development and quality control, writing methods. R.O., laboratory process management, quality control, data collection, writing methods. L.R. and C.W.B.: laboratory processing and wastwater data collection, writing methods. L.T. literature review and initial model evaluation. L.T. and M.S., partner engagement and project management. Y.E.G. and A.S., literature review, paper writing and editing. K.S. and C.N., wastewater monitoring project supervision. Pollock B., Healthy Davis Together project PI, paper editing/revision. M.N., model development, research oversight/supervision, paper editing/revision. H.N.B., Healthy Central Valley Together project PI; project conception, funding, research oversight and collaborator coordination, paper editing/revision.

This research was supported by the Healthy Central Valley Together (HCVT) and Healthy Davis Together (HDT) programs at the University of California, Davis. This research was also supported in part by the NIH Rapid Acceleration of Diagnostics (RADxSM) initiative with federal funds from the National Institute of Biomedical Imaging and Bioengineering, National Institutes of Health. The current contract is funded from the Public Health and Social Services Emergency Fund through the Biomedical Advanced Research and Development Authority, HHS Office of the Assistant Secretary for Preparedness and Response, Department of Health and Human Services, under Contract No. 75N92021C00012. All authors reviewed and approved the final manuscript.

## Supplementary Material

### Results for City of Woodland

Figure S1A illustrates data from Woodland that was used to reconstruct cases from WW. Figure S1B described the smoothed cases and N/PMMoV signals used to provide prediction of cases. Figure S1C describes the training periods used for the analysis; one with adequate testing (shaded in green, denoted by *T*_*A*_) and one in which testing was not adequate (shaded in red, denoted by *T*_*NA*_).

**Figure S1:**
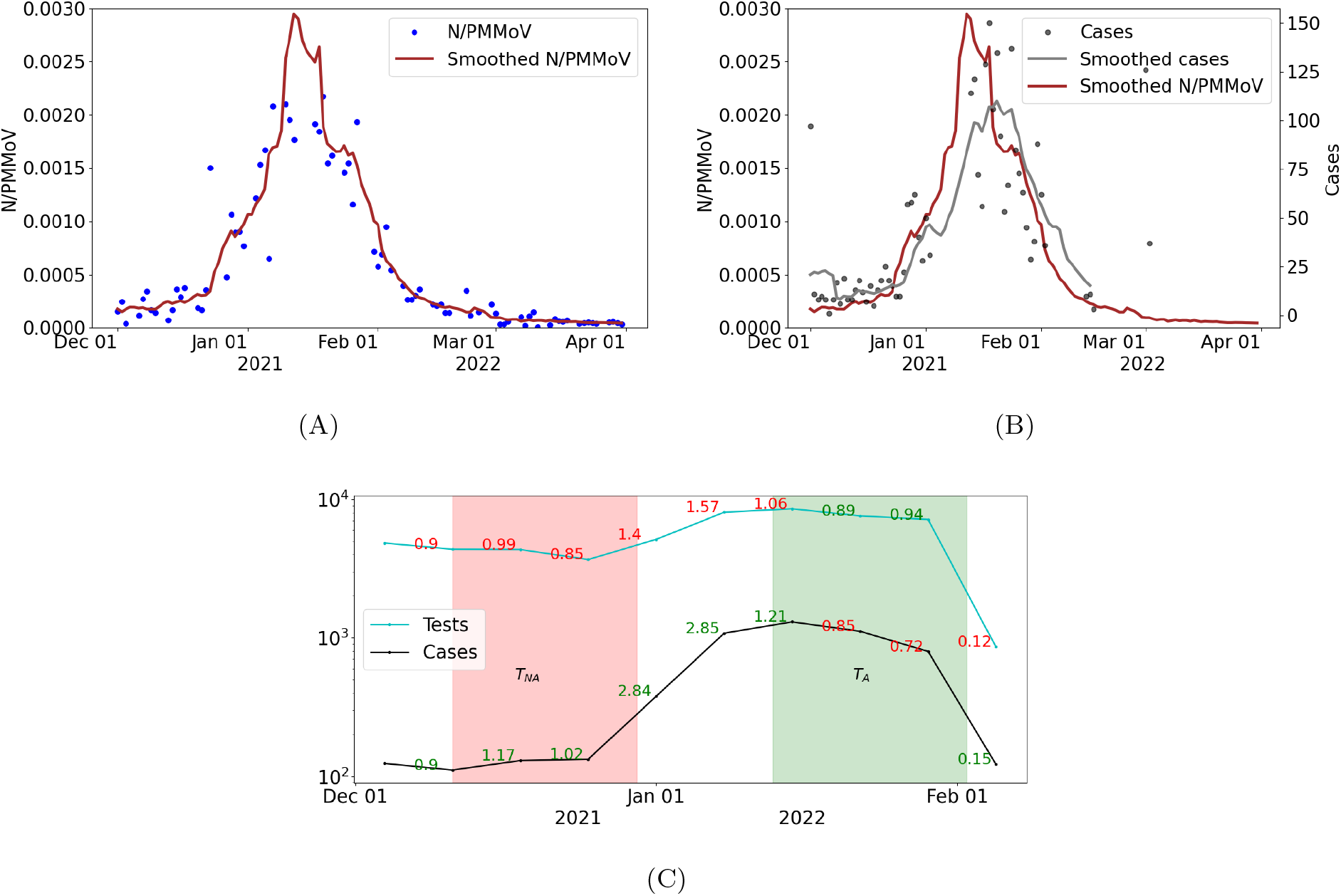
City of Woodland: (A) Wastewater data (N/PMMoV) and 10-day trimmed average (Smoothed N/PMMov) from December 1, 2021 to March 31, 2022; (B) COVID-19 cases, 7-day moving average (Smoothed cases) for cases, and 10-day trimmed average for WW data (Smoothed N/PMMov); (C) Number of tests administered and cases by week, on a log-scale, from December 1, 2021 to February 1, 2022. The week-to-week rate of change in cases and tests are printed with green labels (shaded-region) corresponding to period of adequate testing and red labels (shaded region) for periods in which testing is not adequate.

**Figure S2:**
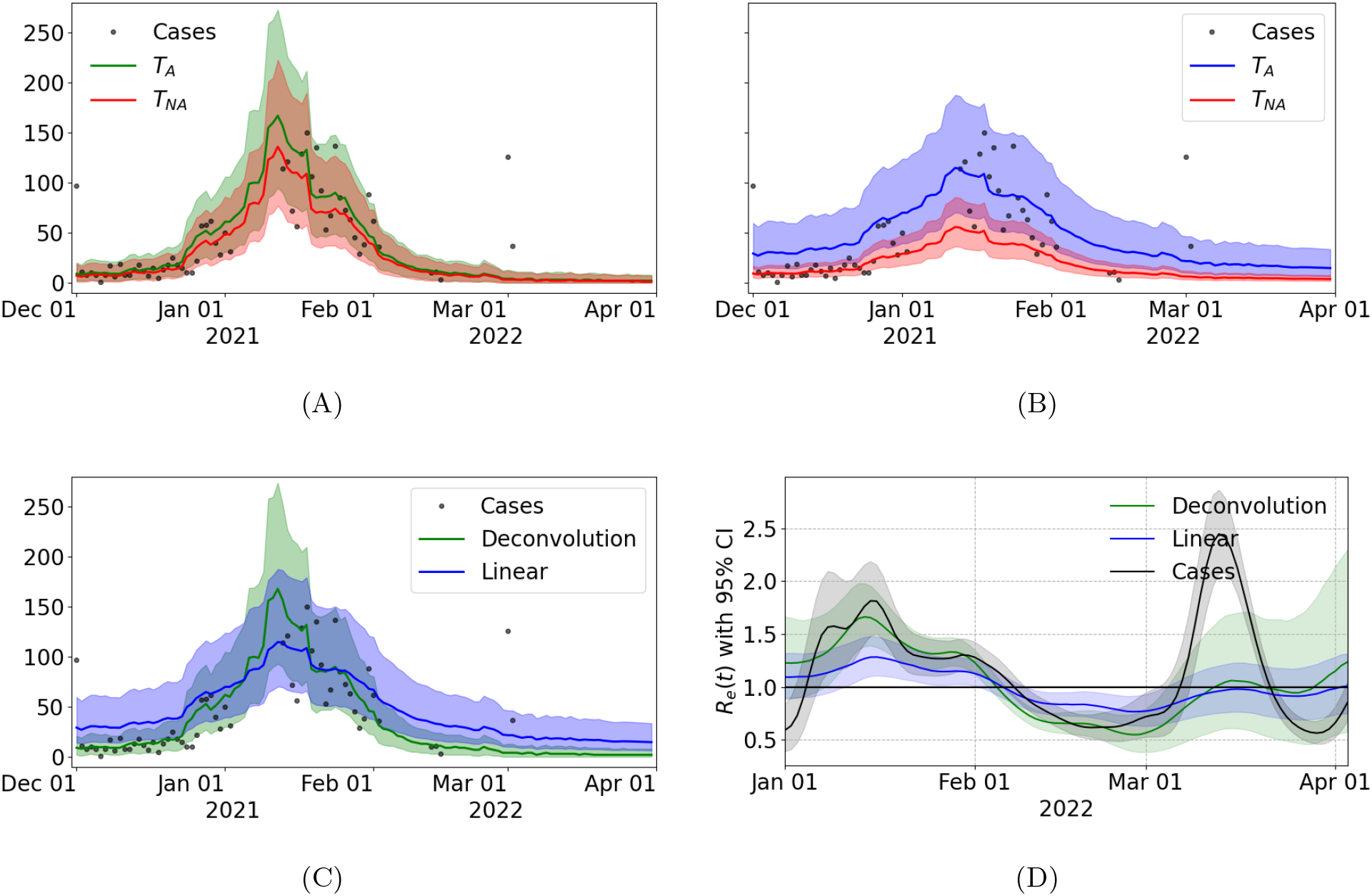
City of Woodland. Reconstruction of incident cases from the deconvolution model and (B) linear regression model using WW data from December 1, 2021 to March 31, 2022. The estimated cases are display (*T*_*A*_: January 13 - February 2, 2022), and red when the models are trained on the period classified as inadequate (*T*_*NA*_: December 11-30, 2021). Solid lines represent median estimates of cases, and 95% prediction intervals are depicted in shaded regions. (C) Predicted cases using the linear regression model (blue) and the deconvolution (green) trained in the period selected as appropriate (*T*_*A*_). (D) Calculation of *R*_*e*_ from case data (black) and from cases reconstructed from WW data using the deconvolution model (green) and the linear regression (blue).

### Results for the UC Davis Campus

Figure S3A illustrates data from UC Davis campus that was used to reconstruct cases from WW. Figure S3B described the smoothed cases and N/PMMoV signals used to provide prediction of cases. Figure S3C describes the training periods used for the analysis; one with adequate testing (shaded in green, denoted by *T*_*A*_) and one in which testing was not adequate (shaded in red, denoted by *T*_*NA*_).

**Figure S3:**
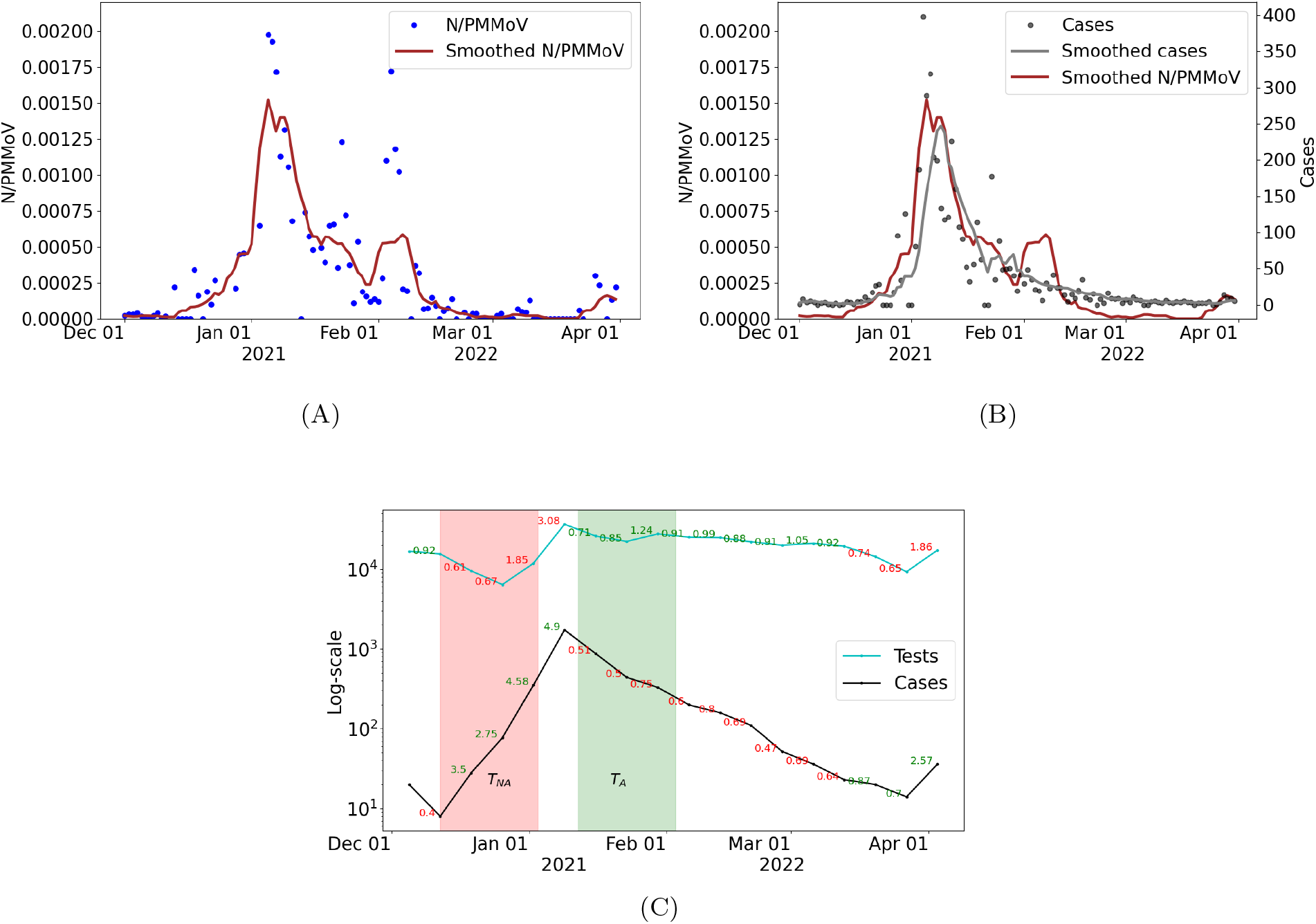
UC Davis Campus. (A) Wastewater data (N/PMMoV) and 10-day trimmed average (Smoothed N/PMMov) from December 1, 2021 to March 31, 2022. (B) COVID-19 cases, 7-day moving average (Smoothed cases) for cases, and 10-day trimmed average for WW data (Smoothed N/PMMov). (C) Number of tests administered and cases by week, on a log-scale, from December 1, 2021 to February 1, 2022. The week-to-week rate of change in cases and tests are printed, with green labels (and shaded-region) corresponding to period of adequate testing and red labels (and shaded region) for inadequate testing.

**Figure S4:**
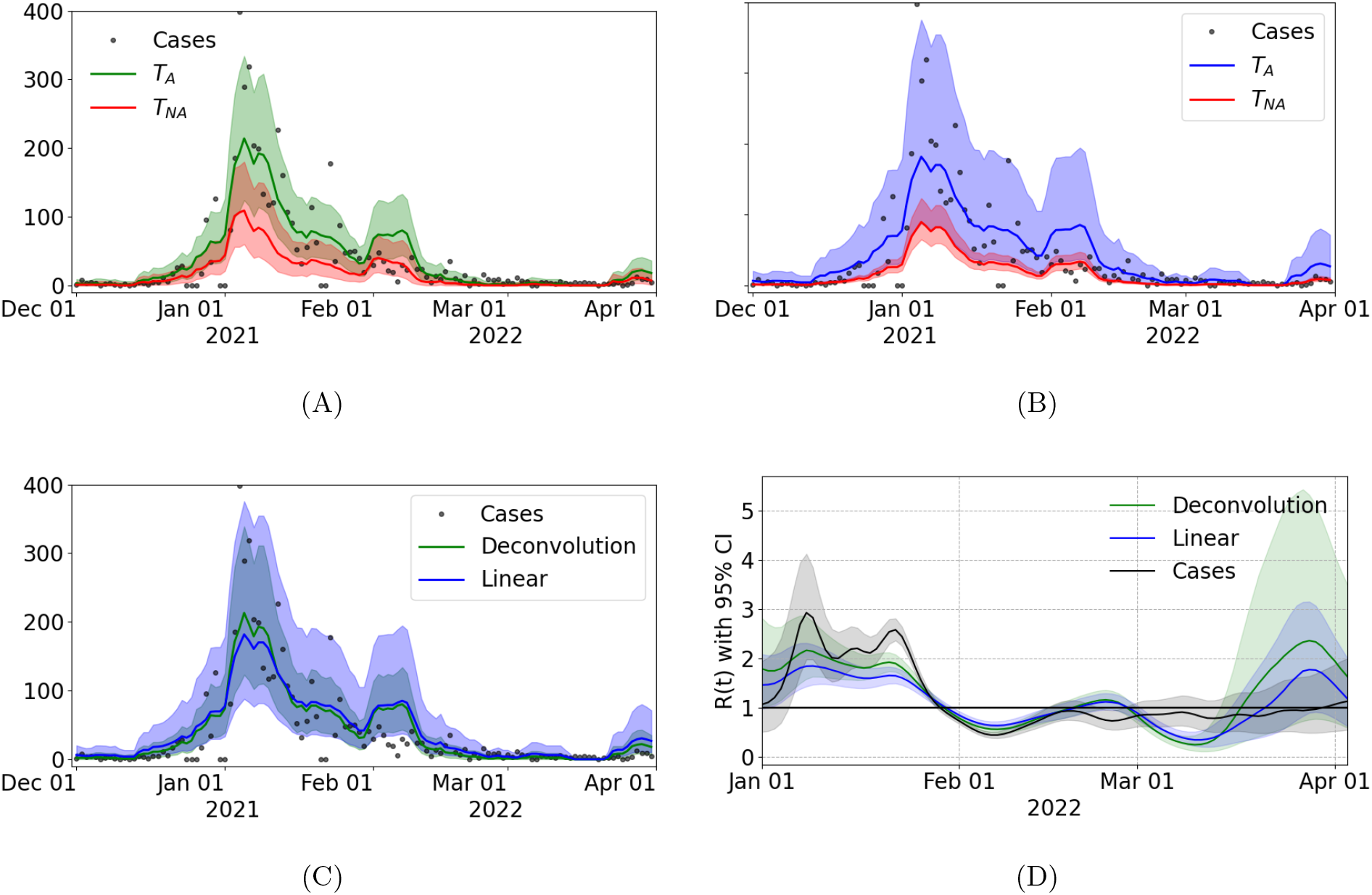
UC Davis Campus. Reconstruction of incident cases from the deconvolution (A) and (B) linear regression models from WW data between December 1, 2021, and March 31, 2022. The estimated cases are display (*T*_*A*_: January 12 - February 3, 2022), and red when the models are trained on the period classified as inadequate (*T*_*NA*_: December 12, 2021 - January 3, 2022). Solid lines represent median estimates of cases, and 95% prediction intervals are depicted in shaded regions. (C) Predicted cases using the linear regression model (blue) and the deconvolution (green) trained in the period selected as appropriate (*T*_*A*_). (d) Effective Re computed with the median of the cases estimated for the deconvolution model (blue) and the linear regression (green).

**Figure S5:**
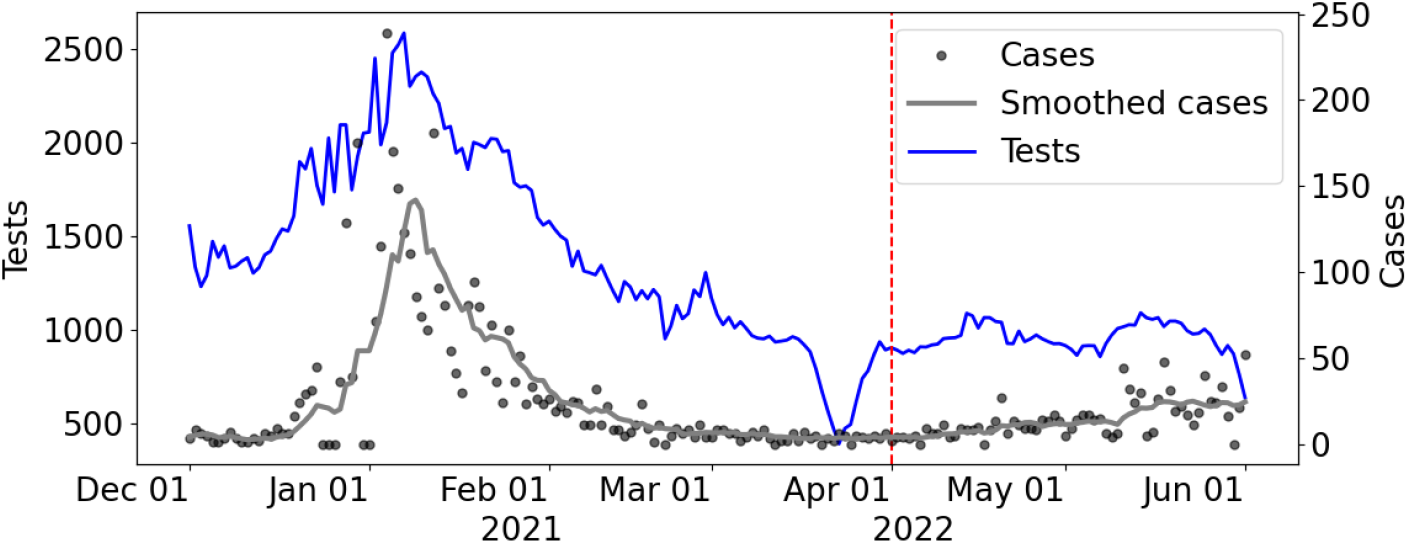
City of Davis data. COVID-19 cases, 7-day moving average (Smoothed cases) for cases, and number of tests administered from December 1, 2021 to June 1, 2022.

### Laboratory quality control and data processing

The sensitivity of the analytical assay was assessed by determining a limit of detection (LOD) and a limit of blank (LOB) following protocols recommended by the ddPCR manufacturer (Bio-Rad Laboratories, 2020, A practical guide for evaluating detection capability using ddPCR.). Fifteen wastewater samples that were initially screened as negative for SARS-CoV-2 in routine wastewater ddPCR monitoring (i.e., extracts had less than 4 positive droplets in merged wells from duplicate analysis) were used to determine the lowest detectable concentrations in ostensibly blank wastewater samples. Selection of these extracts provided a conservative approach to determining the LOB. The selected extracts were re-analyzed by ddPCR to obtain data for four additional replicates for each sample. A non-parametric (rank order) method was then used to select the LOB, since results from the blank were not normally distributed. The ddPCR number of droplets from individual wells were tabulated from lowest to highest. The LOB was set at the value of the concentration measurement for the rank position corresponding to the 95th percentile, calculated as follows: *Rank* = 0.5 + 0.95∗(number of measurements). Since the calculated rank position was a non-integer value, the rank position was rounded up to provide a more conservative LOB. The theoretical LOD was set as LOB plus two times the standard deviation of all replicate results (Biorad, 2020). The LOD and LOB are reported in Table S5. In terms of droplet numbers in the blank samples, the highest numbers of positive droplets in the merged wells (four replicates) amongst the fifteen blank samples were 6 (N1) and 8 (N2). Since routine wastewater samples were analyzed in duplicate, 3 (N1) and 4 (N2) droplets were set as the cutoff to mark samples below the droplet threshold. Samples were also considered below the droplet threshold if there were fewer N1 and N2 droplets twice the number of droplets in the extraction control blank analyzed on the same day. Runs with an extraction control blank that had > 15 positive droplets in either N1 or N2 were considered contaminated and extracts were re-processed.

If samples passed all checks, the relative concentration of N gene was calculated as follows. Duplicate results for each target were merged, and the concentration of each target in the ddPCR reaction was calculated assuming a Poisson distribution using the QXOne Software 1.1.1 Standard Addition (Bio-Rad). The average SARS-CoV-2 RNA concentration in the initial wastewater sample was calculated from the average of the N1 and N2, corrected for sample and reagent volumes used, and reported as genome copies (gc) per mL wastewater. BCoV was detected in 100% of spiked samples, and concentrations of targets were not corrected for BCoV recovery efficiency. The average SARS-CoV-2 RNA concentration (*N*), was divided by the concentration of PMMoV. If N1 or N2 merged droplet counts were below the minimum droplet threshold, the target was excluded from the average concentration. If both N1 and N2 targets were below the droplet threshold, the concentration was reported as 0.

**Table S1:**
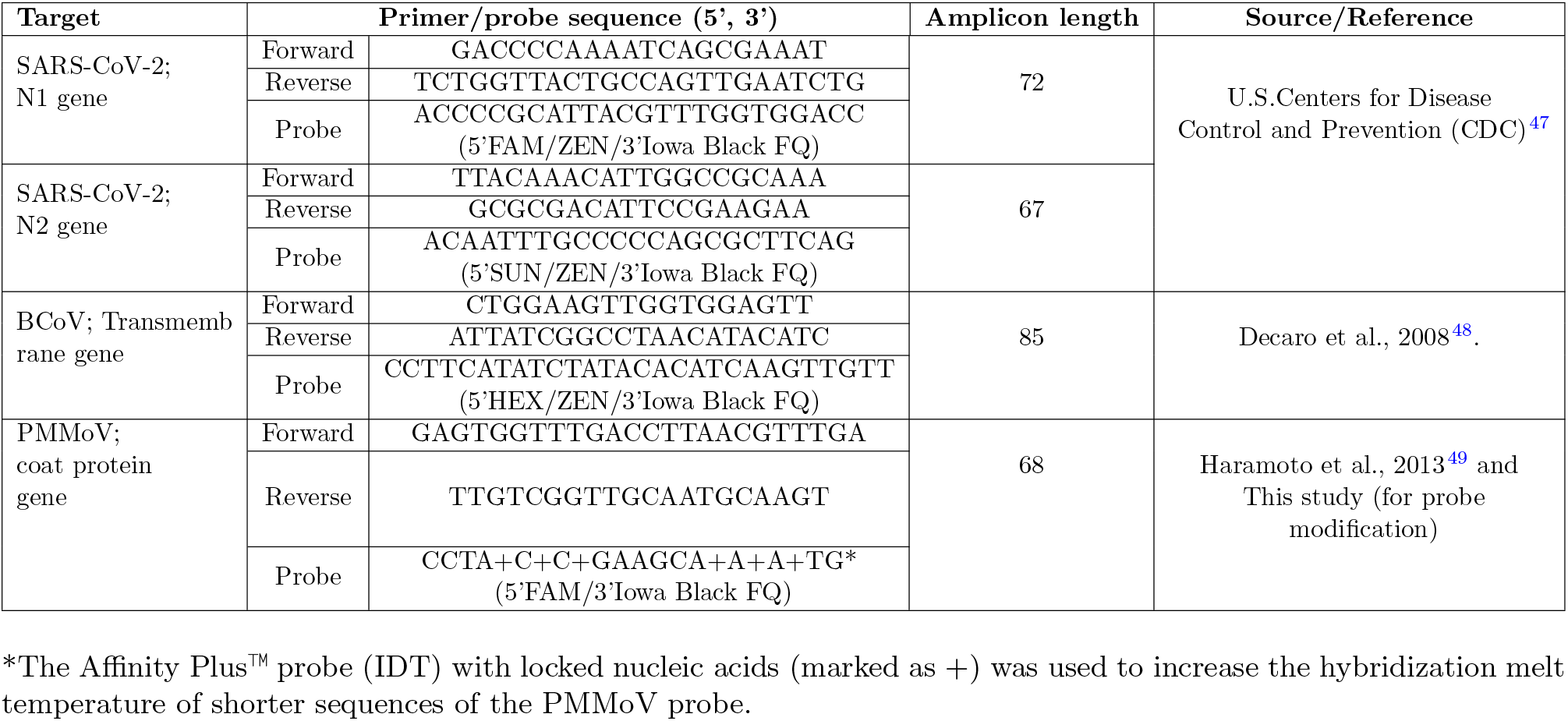
RT-ddPCR primers and probes used in this study.

**Table S2:**
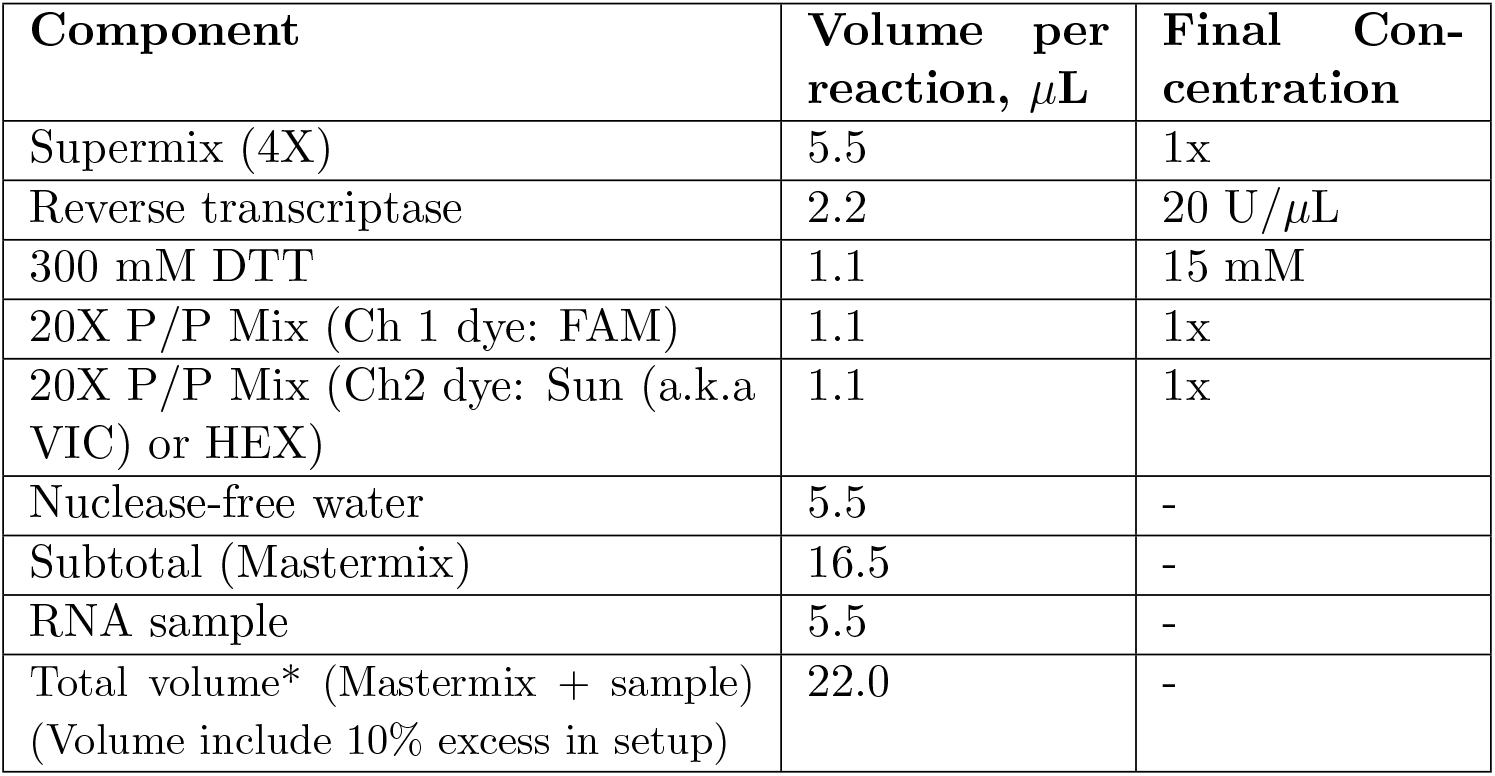
Preparation of the duplex One-Step RT-ddPCR reaction.

| Component | Volume per reaction, $\mu\text{L}$ | Final Concentration |
| --- | --- | --- |
| Supermix (4X) | 5.5 | 1x |
| Reverse transcriptase | 2.2 | 20 U/$\mu\text{L}$ |
| 300 mM DTT | 1.1 | 15 mM |
| 20X P/P Mix (Ch 1 dye: FAM) | 1.1 | 1x |
| 20X P/P Mix (Ch2 dye: Sun (a.k.a VIC) or HEX) | 1.1 | 1x |
| Nuclease-free water | 5.5 | - |
| Subtotal (Mastermix) | 16.5 | - |
| RNA sample | 5.5 | - |
| Total volume* (Mastermix + sample)<br>(Volume include 10% excess in setup) | 22.0 | - |

**Table S3:** Preparation of 20X primer/probe Mix (p/p Mix).

| Target | Recipe |  |  |  |
| --- | --- | --- | --- | --- |
| | Reagent | Initial concentration | Volume added ($\mu\text{L}$) | Final concentration in ddPCR reaction |
| 20X p/p Mix | Forward | 100 $\mu\text{M}$ | 45.0 | 900 nM |
| | Reverse | 100 $\mu\text{M}$ | 45.0 | 900 nM |
| | Probe | 100 $\mu\text{M}$ | 12.5 | 250 nM |
|  | nuclease-free water | - | 147.5 | - |
|  | Total volume |  | 250 |  |

**Table S4:**
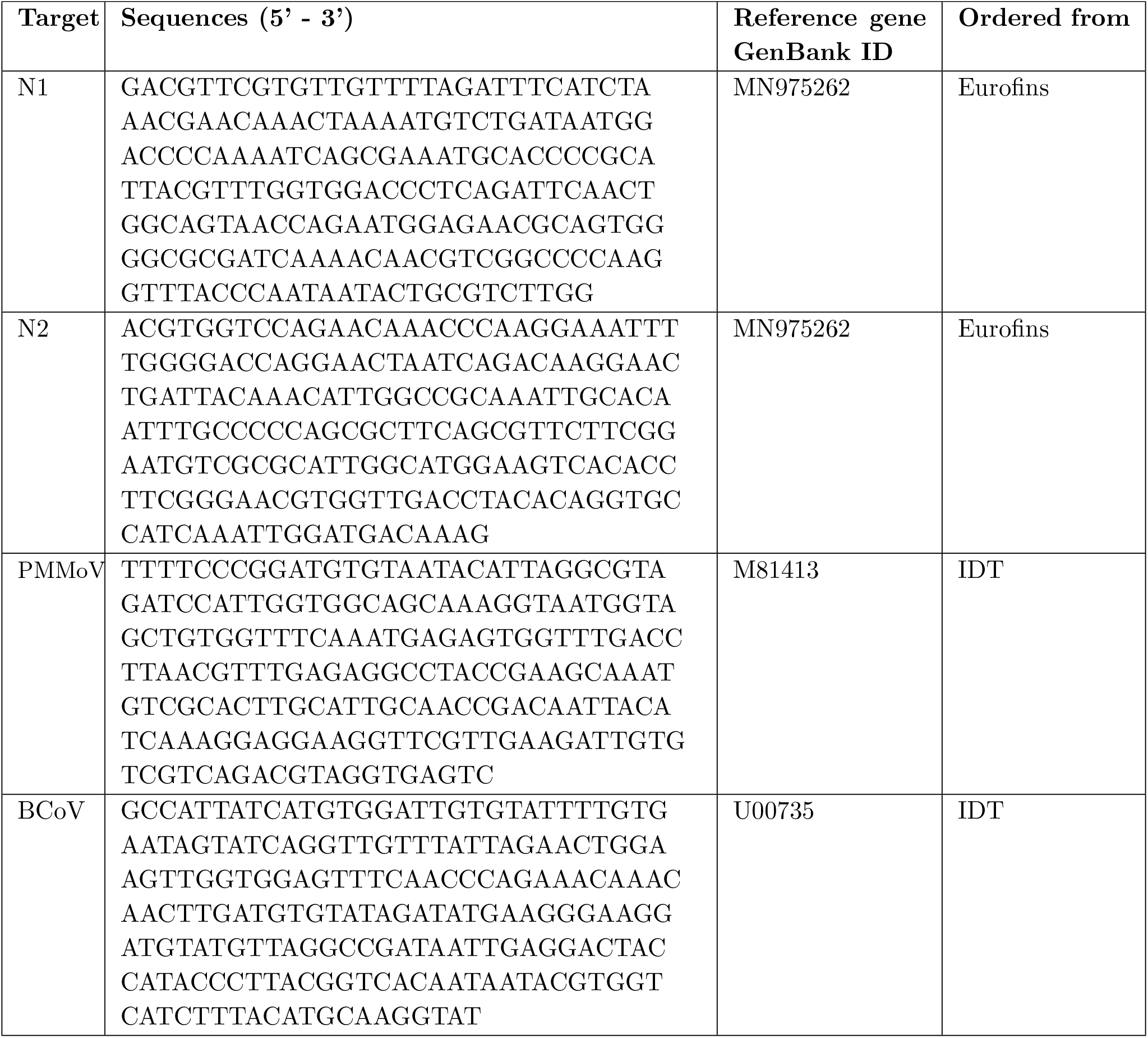
Synthesized gene fragments used for positive controls in ddPCR.

**Table S5:**
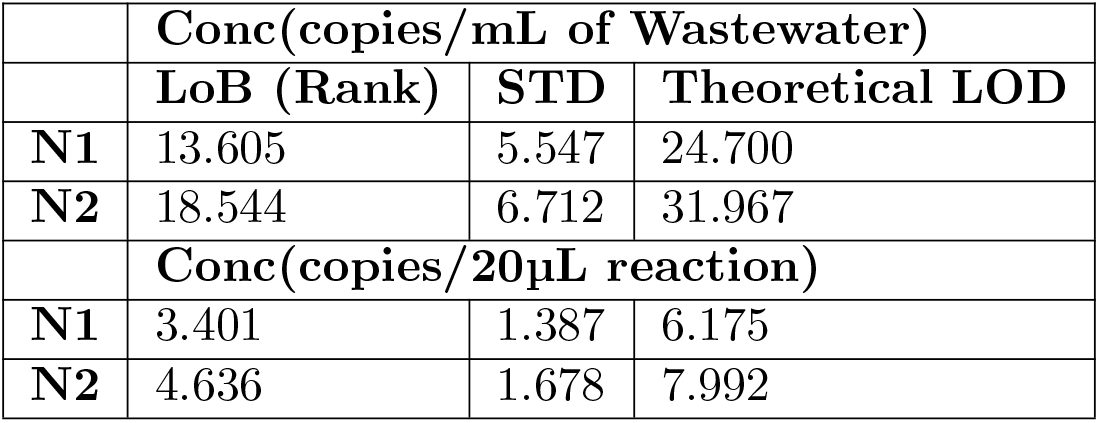
Limit of Blank and Limit of Detection.

|  | Conc(copies/mL of Wastewater) |  |  |
| --- | --- | --- | --- |
|  | LoB (Rank) | STD | Theoretical LOD |
| N1 | 13.605 | 5.547 | 24.700 |
| N2 | 18.544 | 6.712 | 31.967 |
|  | Conc(copies/20µL reaction) |  |  |
| N1 | 3.401 | 1.387 | 6.175 |
| N2 | 4.636 | 1.678 | 7.992 |

**Table S6:**
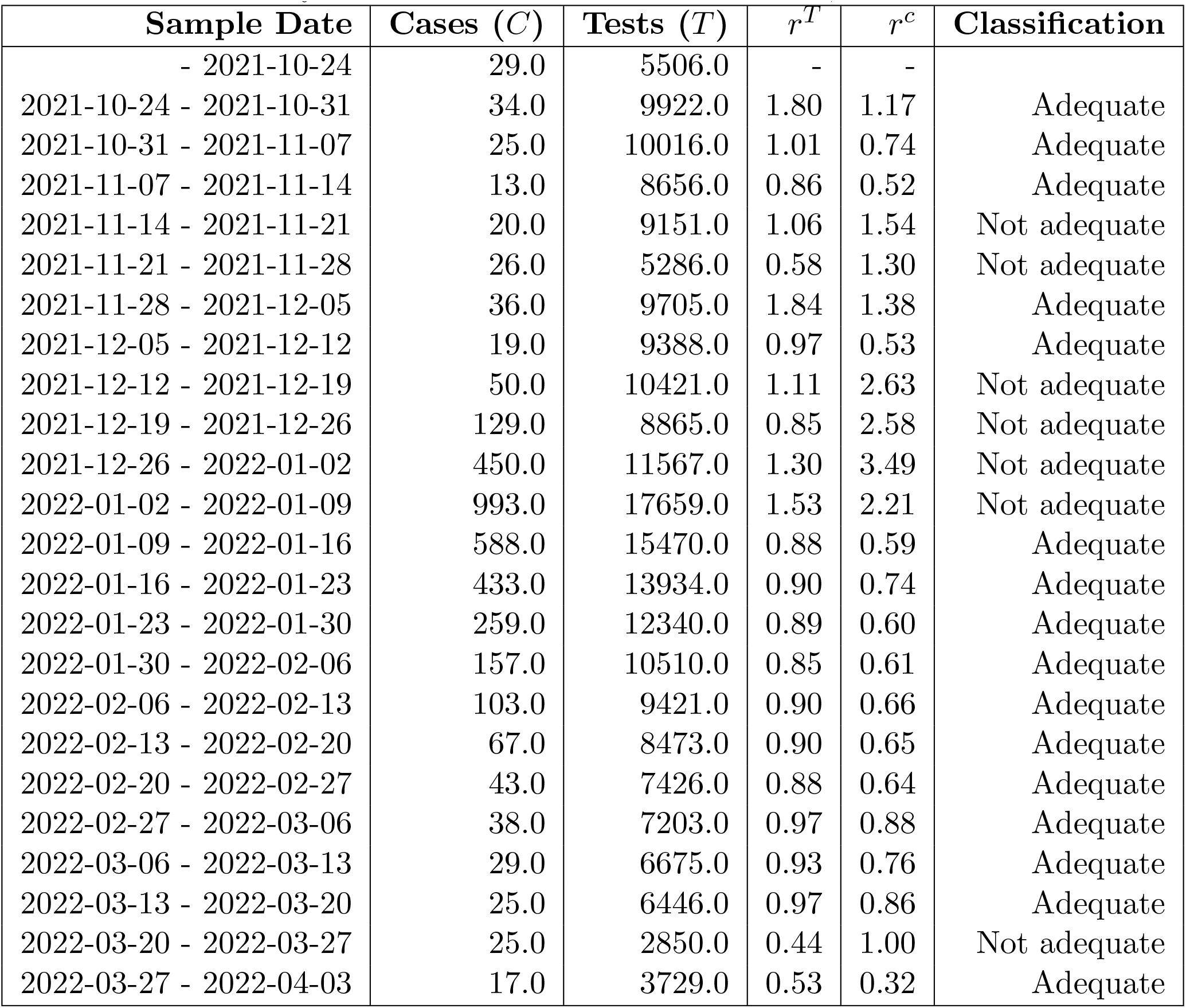
Weekly cases for Davis between October 21, 2021 and March 31.

| Sample Date | Cases ($C$) | Tests ($T$) | $r^T$ | $r^C$ | Classification |
| --- | --- | --- | --- | --- | --- |
| - 2021-10-24 | 29.0 | 5506.0 | - | - |  |
| 2021-10-24 - 2021-10-31 | 34.0 | 9922.0 | 1.80 | 1.17 | Adequate |
| 2021-10-31 - 2021-11-07 | 25.0 | 10016.0 | 1.01 | 0.74 | Adequate |
| 2021-11-07 - 2021-11-14 | 13.0 | 8656.0 | 0.86 | 0.52 | Adequate |
| 2021-11-14 - 2021-11-21 | 20.0 | 9151.0 | 1.06 | 1.54 | Not adequate |
| 2021-11-21 - 2021-11-28 | 26.0 | 5286.0 | 0.58 | 1.30 | Not adequate |
| 2021-11-28 - 2021-12-05 | 36.0 | 9705.0 | 1.84 | 1.38 | Adequate |
| 2021-12-05 - 2021-12-12 | 19.0 | 9388.0 | 0.97 | 0.53 | Adequate |
| 2021-12-12 - 2021-12-19 | 50.0 | 10421.0 | 1.11 | 2.63 | Not adequate |
| 2021-12-19 - 2021-12-26 | 129.0 | 8865.0 | 0.85 | 2.58 | Not adequate |
| 2021-12-26 - 2022-01-02 | 450.0 | 11567.0 | 1.30 | 3.49 | Not adequate |
| 2022-01-02 - 2022-01-09 | 993.0 | 17659.0 | 1.53 | 2.21 | Not adequate |
| 2022-01-09 - 2022-01-16 | 588.0 | 15470.0 | 0.88 | 0.59 | Adequate |
| 2022-01-16 - 2022-01-23 | 433.0 | 13934.0 | 0.90 | 0.74 | Adequate |
| 2022-01-23 - 2022-01-30 | 259.0 | 12340.0 | 0.89 | 0.60 | Adequate |
| 2022-01-30 - 2022-02-06 | 157.0 | 10510.0 | 0.85 | 0.61 | Adequate |
| 2022-02-06 - 2022-02-13 | 103.0 | 9421.0 | 0.90 | 0.66 | Adequate |
| 2022-02-13 - 2022-02-20 | 67.0 | 8473.0 | 0.90 | 0.65 | Adequate |
| 2022-02-20 - 2022-02-27 | 43.0 | 7426.0 | 0.88 | 0.64 | Adequate |
| 2022-02-27 - 2022-03-06 | 38.0 | 7203.0 | 0.97 | 0.88 | Adequate |
| 2022-03-06 - 2022-03-13 | 29.0 | 6675.0 | 0.93 | 0.76 | Adequate |
| 2022-03-13 - 2022-03-20 | 25.0 | 6446.0 | 0.97 | 0.86 | Adequate |
| 2022-03-20 - 2022-03-27 | 25.0 | 2850.0 | 0.44 | 1.00 | Not adequate |
| 2022-03-27 - 2022-04-03 | 17.0 | 3729.0 | 0.53 | 0.32 | Adequate |

